## Supplementary Figures for "The genetic architecture of human cerebellar morphology supports a key role for the cerebellum in human evolution and psychopathology"

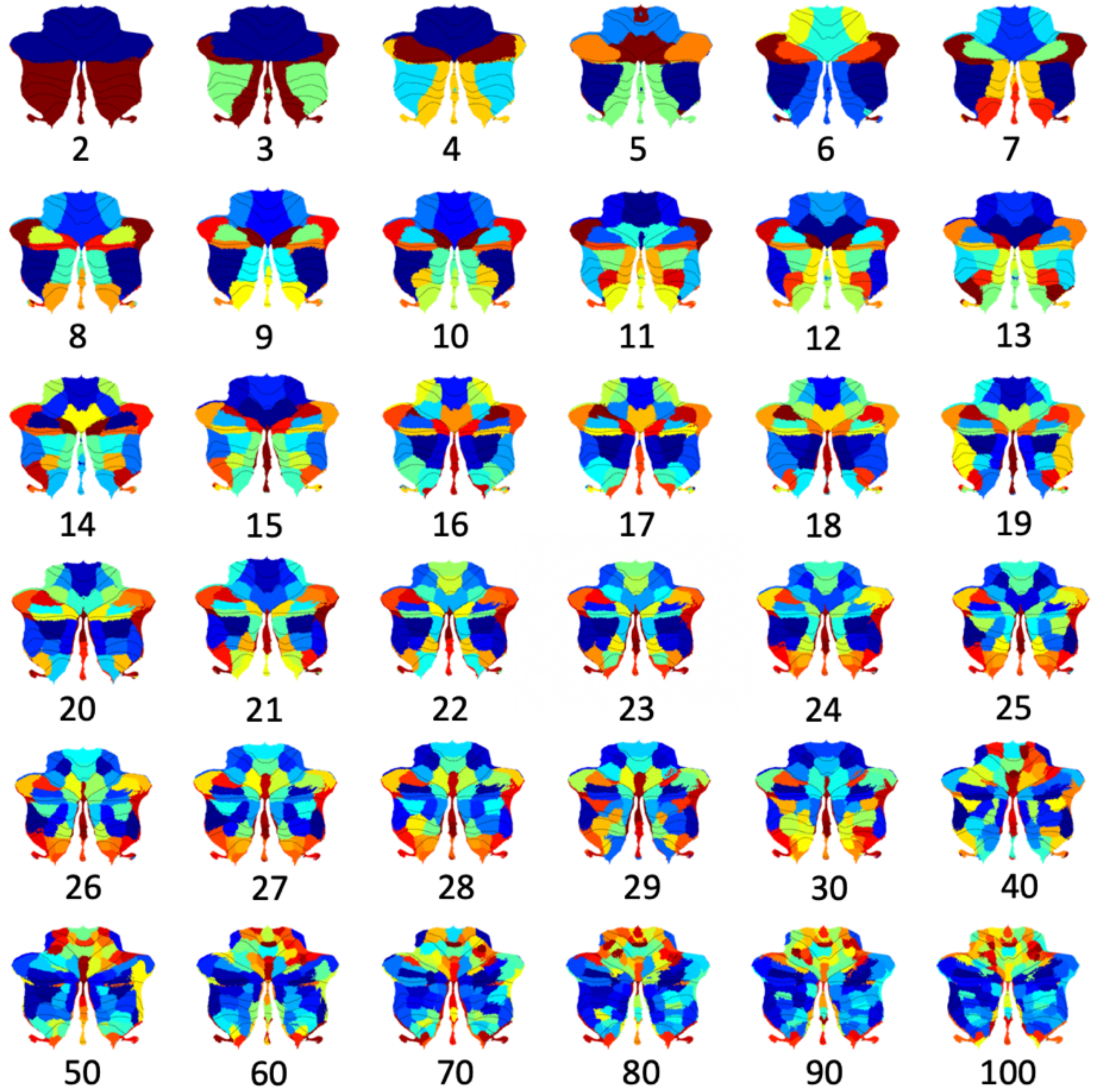


***Supplementary Figure 1:*** *Summary maps of cerebellar components resulting from running non-negative matrix factorization (NNMF) on voxel-wise cerebellar grey matter maps for the full sample (n = 28,212) using all tested model orders. The two-component solution reveals a primary division running along the horizontal fissure. Medial-to-lateral divisions within cerebellar lobules already emerged with a model order of three and remained prominent for all higher model orders.*

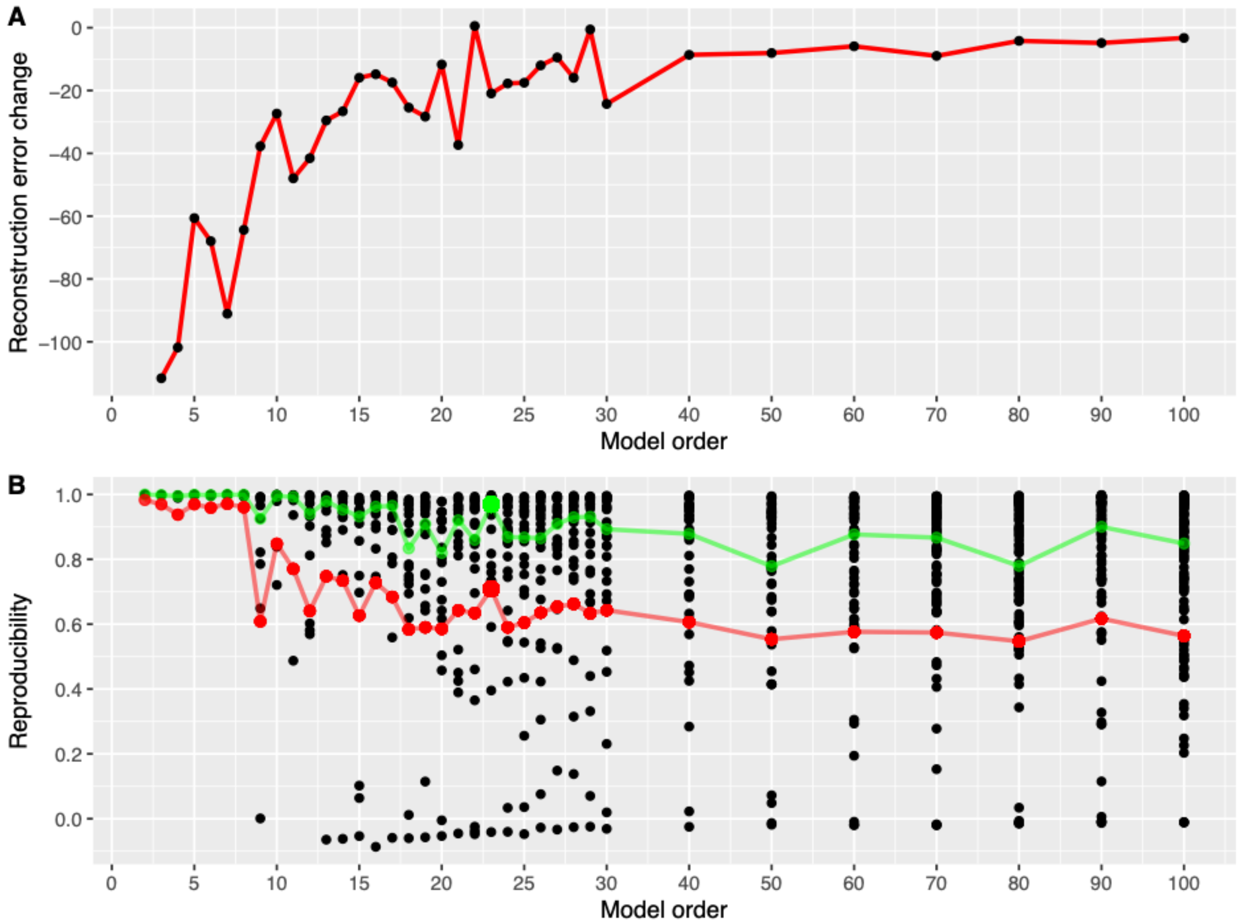


***Supplementary Figure 2: Variance explained and reproducibility as a function of model order. A:*** *Reconstruction error change (indicating increased original data variance explained by the NNMF solution) when moving to higher model orders, plotted from model orders 3 to 100.* ***B:*** *Split-half reproducibility of NNMF component maps as a function of model order. Black dots indicate spatial correlations between matched pairs of components, while the green line shows the median pairwise correlation and the red line shows the adjusted Rand index between binarized (i.e. winner-takes-all) categorical component maps for a given model order.*

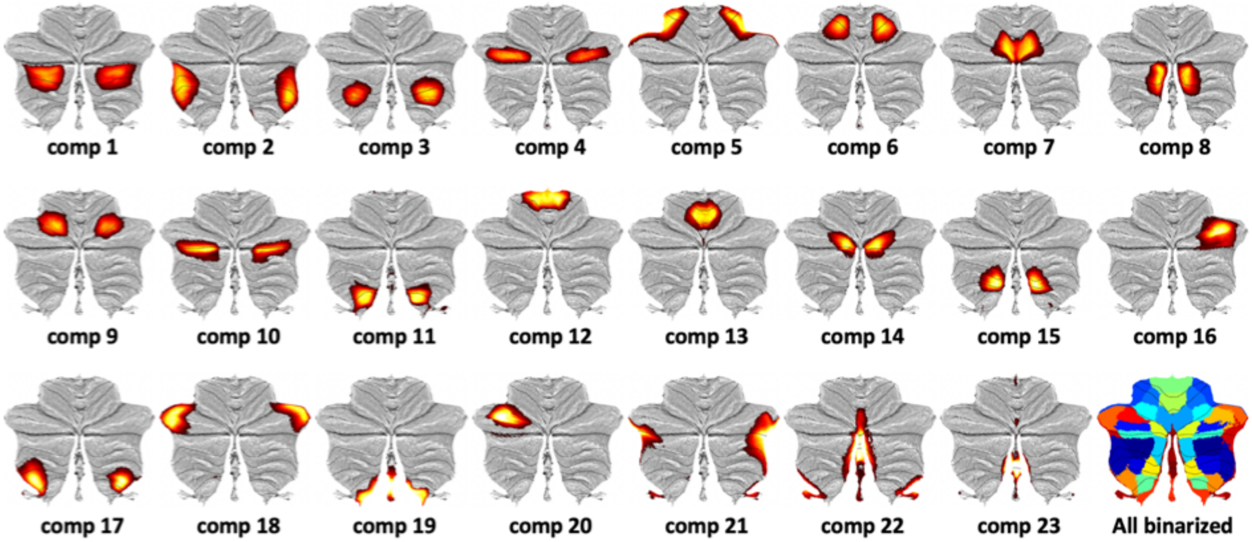


***Supplementary Figure 3:*** *Summary maps of cerebellar components resulting from running non-negative matrix factorization (NNMF) of cerebellar grey matter maps for the full sample (n = 28,212) using the selected model order of 23. As can be seen, NNMF yielded sparce and largely symmetrical/bilateral components. The bottom right plot shows the binarized version of the map, where each voxel is assigned to the component with the highest weight ("winner-tales-all").*

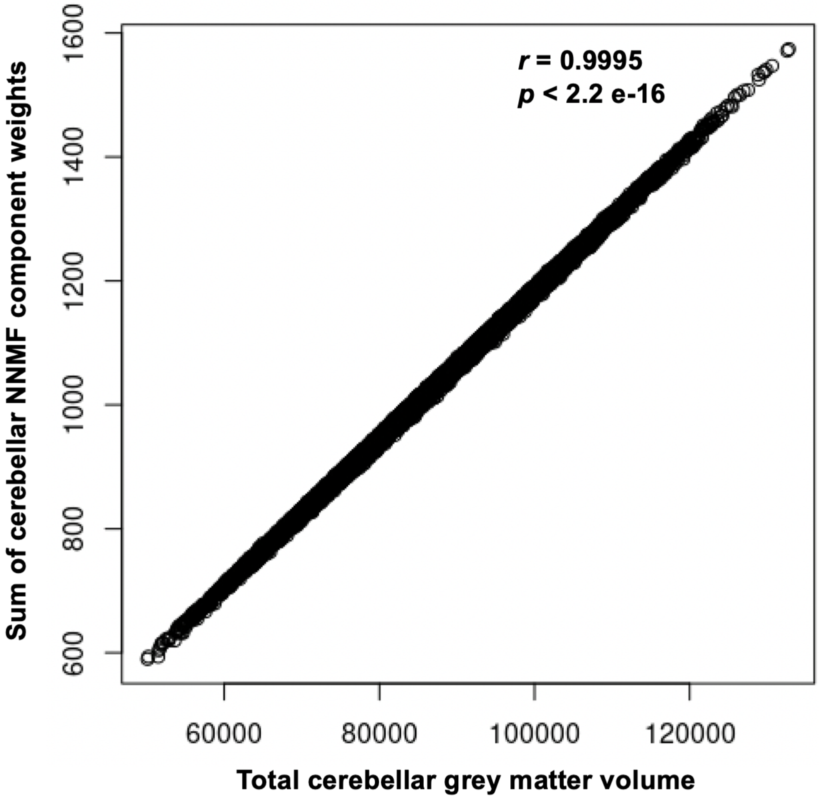


***Supplementary Figure 4: The sum of cerebellar NNMF subject weights correlates tightly with total cerebellar grey matter volume.*** *This demonstrates that the subjects weight resulting from the data-driven decomposion using NNMF (reflecting the degree to which a particular cerebellar structural covariance pattern is expressed) preserves* inter-individual variation in cerebellar volume.

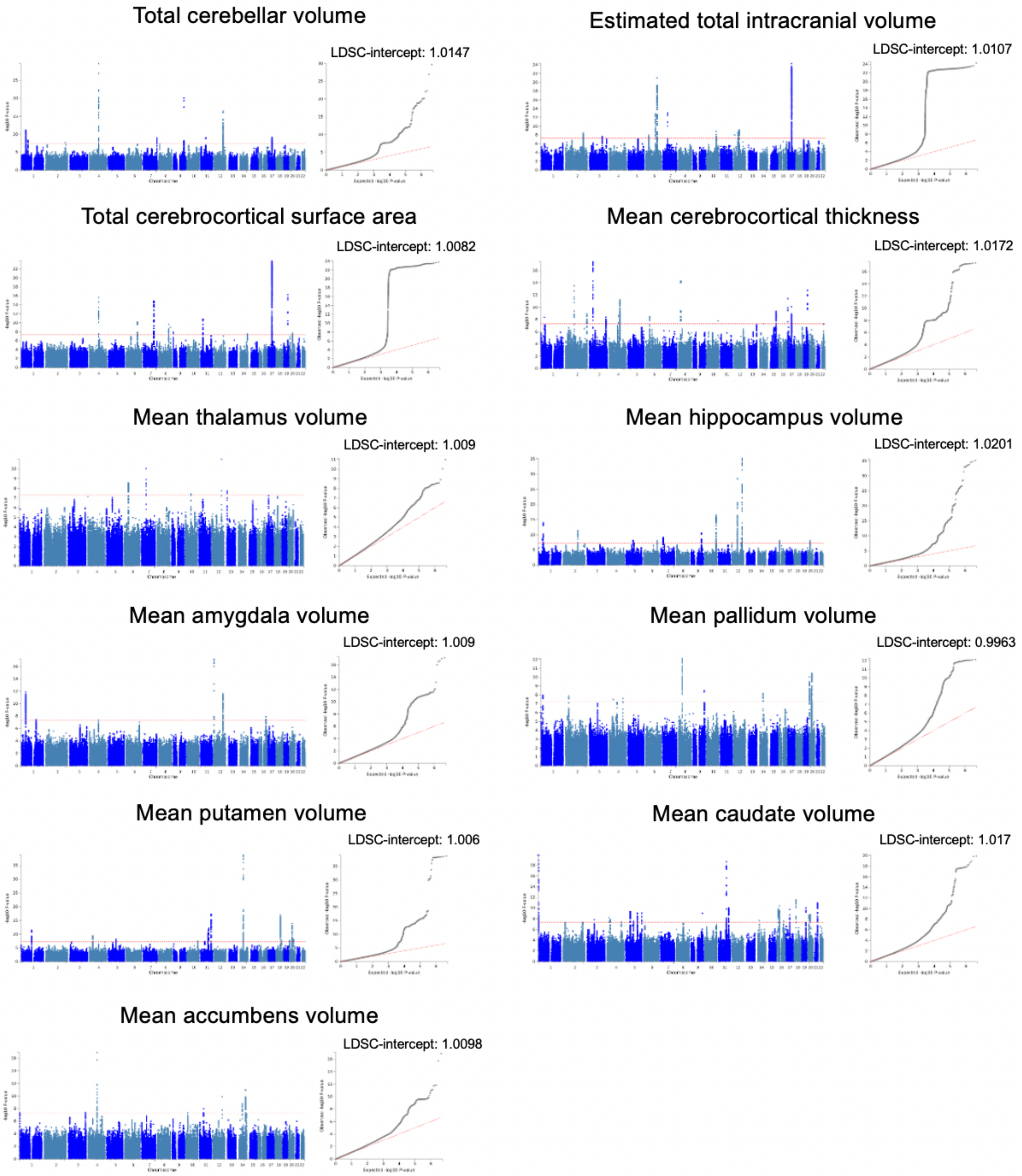


***Supplementary Figure 5:*** *Manhattan and QQ-plots for univariate GWASs on cerebral comparison phenotypes in the current sample. Each plot also provides the intercept from LD-score regression analyses (LDSC-intercept), where values close to 1 indicate no or minimal inflation due to bias (such as cryptic relatedness and population stratification). Manhattan- and QQ-plots from FUMA (https://fuma.ctglab.nl/).*

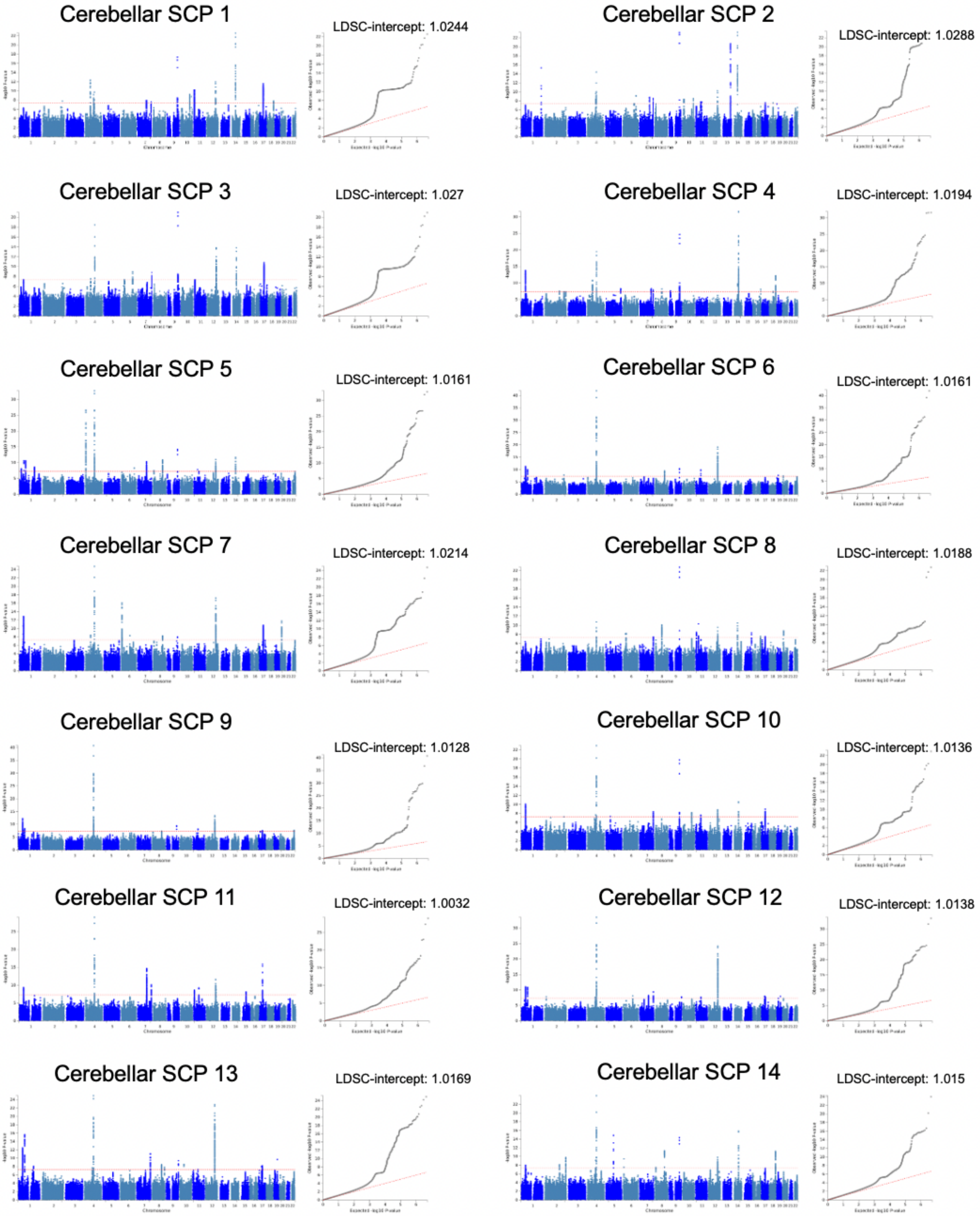


***Supplementary Figure 6:*** *Manhattan and QQ-plots (with LDSC-intercepts) for univariate GWASs on cerebellar structural covariance patterns (SCPs) 1-14. Manhattan- and QQ-plots from FUMA (https://fuma.ctglab.nl/).*

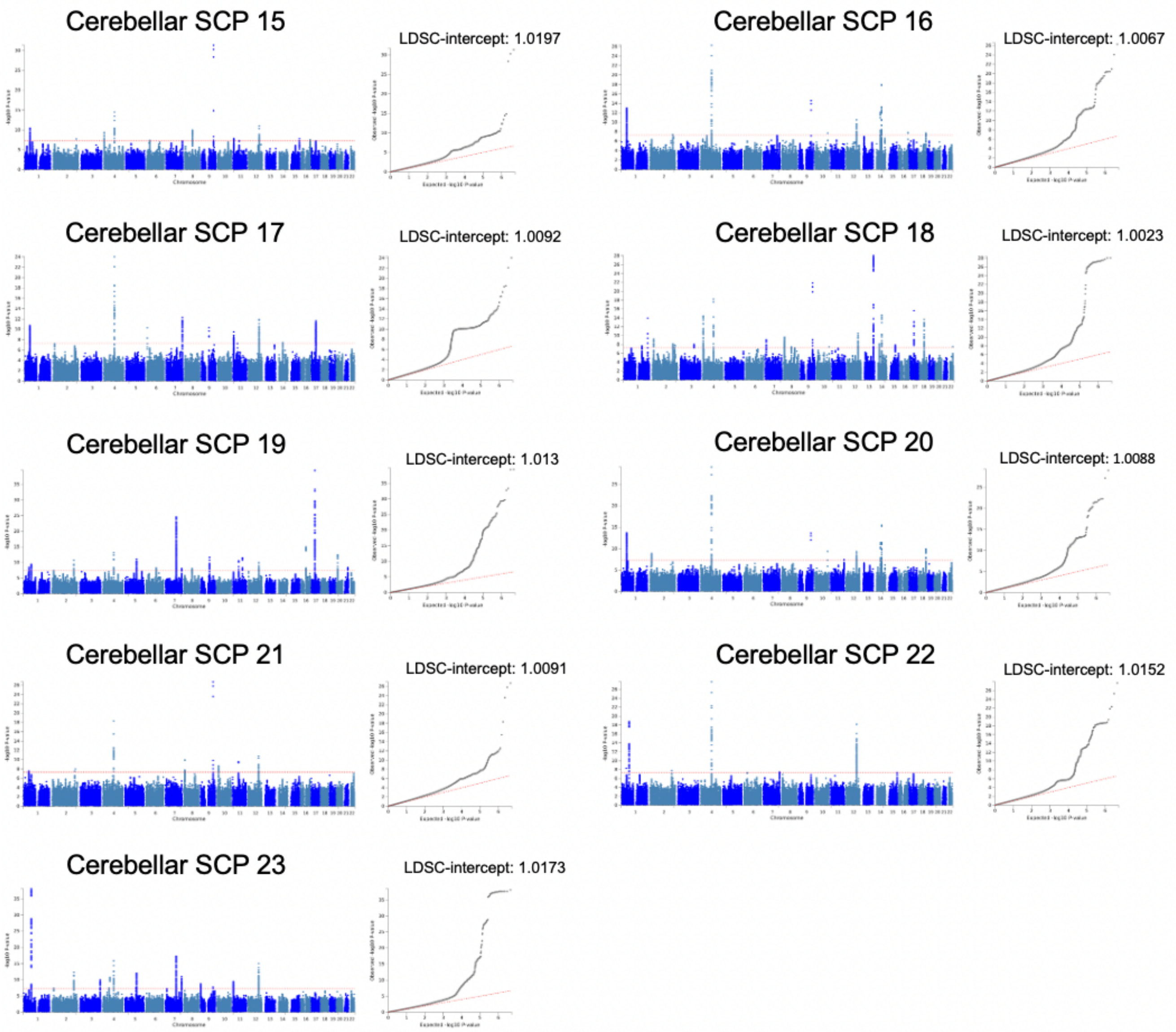


***Supplementary Figure 7:*** *Manhattan and QQ-plots (with LDSC-intercepts) for univariate GWASs on cerebellar structural covariance patterns (SCPs) 15-23. Manhattan- and QQ-plots from FUMA (https://fuma.ctglab.nl/).*

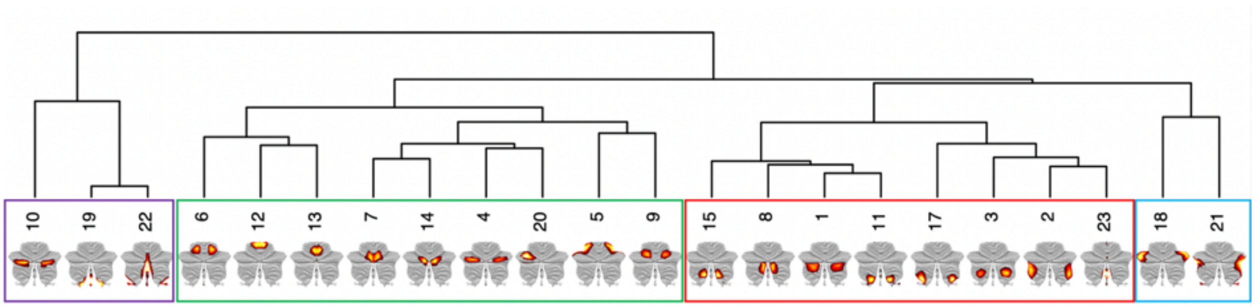


***Supplementary Figure 8:*** *Hierarchical clustering solution for gene expression profiles (derived from the Allen Human Brain Atlas) across 22 of the 23 components (insufficient gene expression data was available for component 16). The clusters in the green and red boxes largely conform to the anterior and posterior clusters evident in the clustering based on genetic correlations between components, while the clusters in the violet and blue boxes were only evident in the gene expression data.*

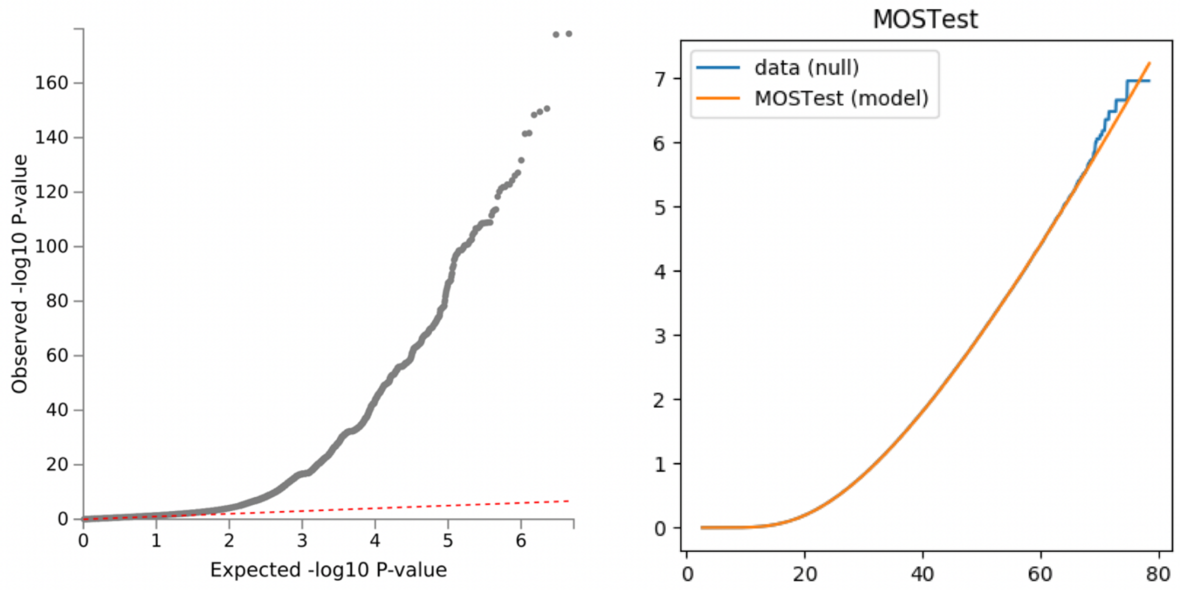


***Supplementary Figure 9:*** *Quantile-quantile plots from MOSTest analysis. The left panel depicts the signal resulting from the MOSTest analysis,and reveals a clear genetic signal with no indication of inflation (LDSC-intercept: 1.0352). The right panel shows test statistics under the null hypothesis (from permuted data): The "Observed" plot shows empirical distribution of the test statistic; "Fitted" plot shows p-values calculated from gamma(a,b) distribution (MOSTest) and Beta(a,b) distribution (min-P) after fitting the two parameters to the observed data. Coincidence of the "Fitted" and "Observed" plots indicate that under null MOSTest p-values are uniformly distributed, as expected for a statistical test with well calibrated type I error.*

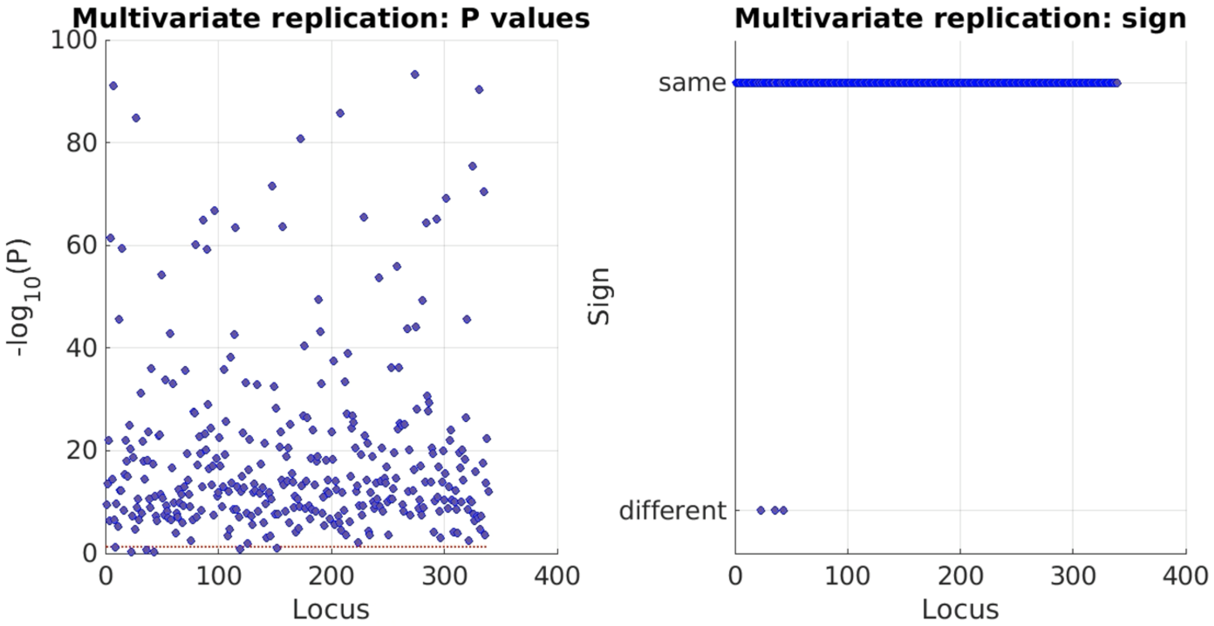


***Supplementary Figure 10:*** *Results from the multivariate replication analysis, where the discovery sample feature z-scores from the 339 locus lead SNPs present in both samples were multiplied with raw data from the replication sample, yielding one composite multivariate score for each participant. For each lead SNP, these replication sample composite scores were correlated with the replication sample genetic data. The left panel shows the resulting -log10 p-values from this multivariate replication test, with the red horizontal line denoting a nominal significance threshold of p<0.05. The right panel shows results from the sign test, which revealed that 99.4% of SNPs showed the same direction of effects across discovery and replication samples.*

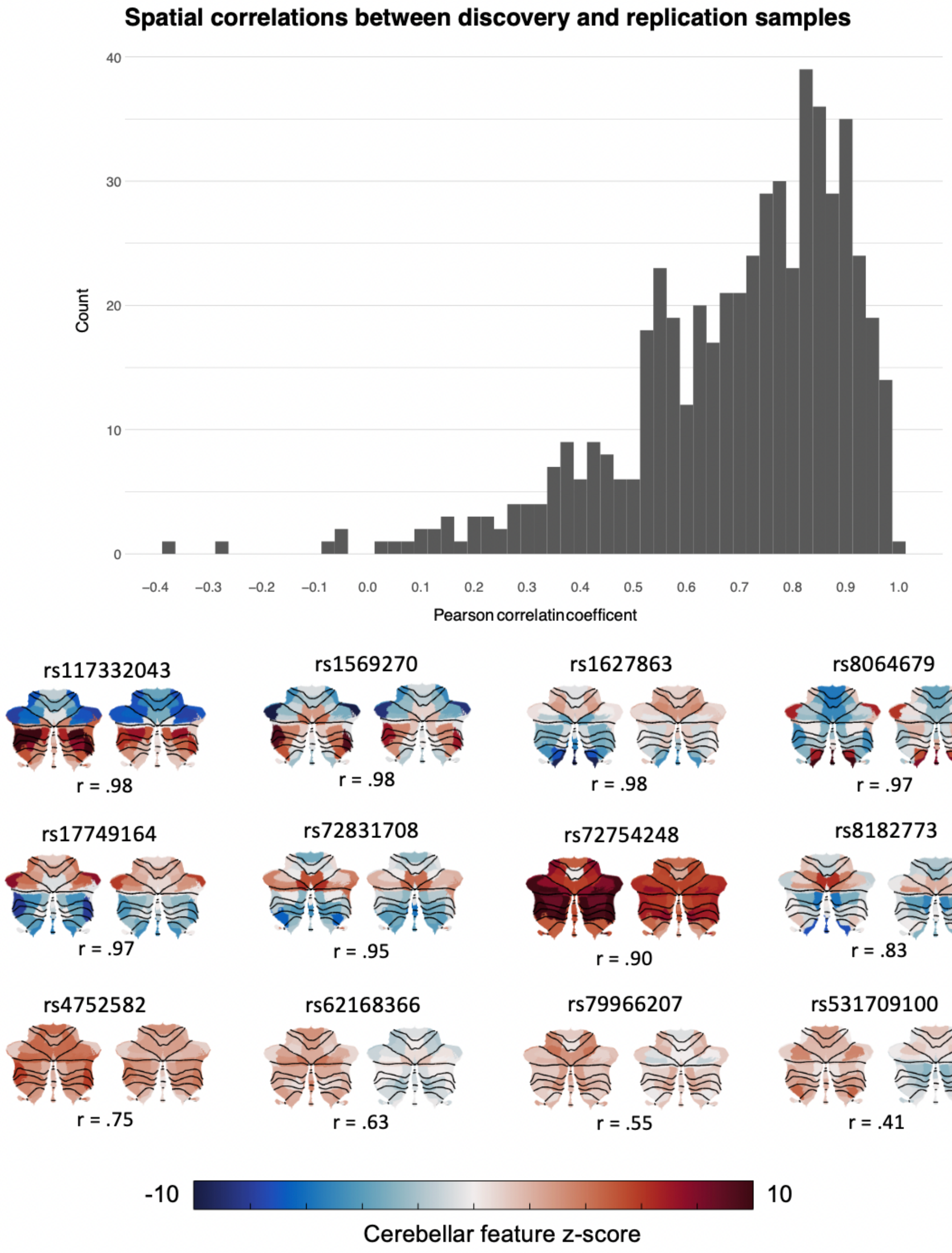


***Supplementary Figure 11: Top panel:*** *Distribution of Pearson correlation coefficients between 341 discovery sample loci lead SNP component z-scores and component z-scores resulting from an independent MOSTest analysis performed on the same SNPs in the replication sample.* ***Bottom panel:*** *Discovery (right) and replication (left) sample Z-scores projected onto the cerebellar cortex for a few selected SNPs, with the Pearson correlation coefficient given below each pair.*

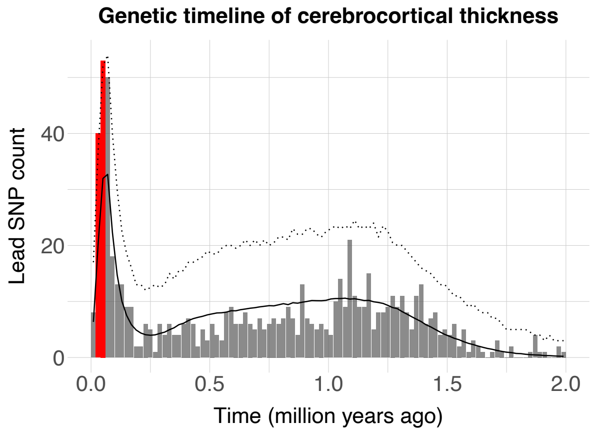


***Supplementary Figure 12: Figure 4: Lead SNPs associated with cerebrocortical thickness are enriched for evolutionary recent mutations in the human genome:*** *Histogram of estimated SNP age (ranging from 0 to 2 million years, in bins on 20,000 years) for 923 independent lead SNPS associated with regional cerebrocortical thickness. The solid black and dotted lines denote the mean and upper 95th confidence interval (Bonferroni corrected across 100 time-bins) derived from a null model constructed from 10,000 of equally sized sets of SNPs randomly drawn from the Human Genome Dating Atlas of Variant Age (after matching these to cerebrocortical thickness-related SNPs in terms of minor allele frequencies). Red bars denote time-bins of significant positive enrichment. See Supplementary Data 12 for full numerical results.*

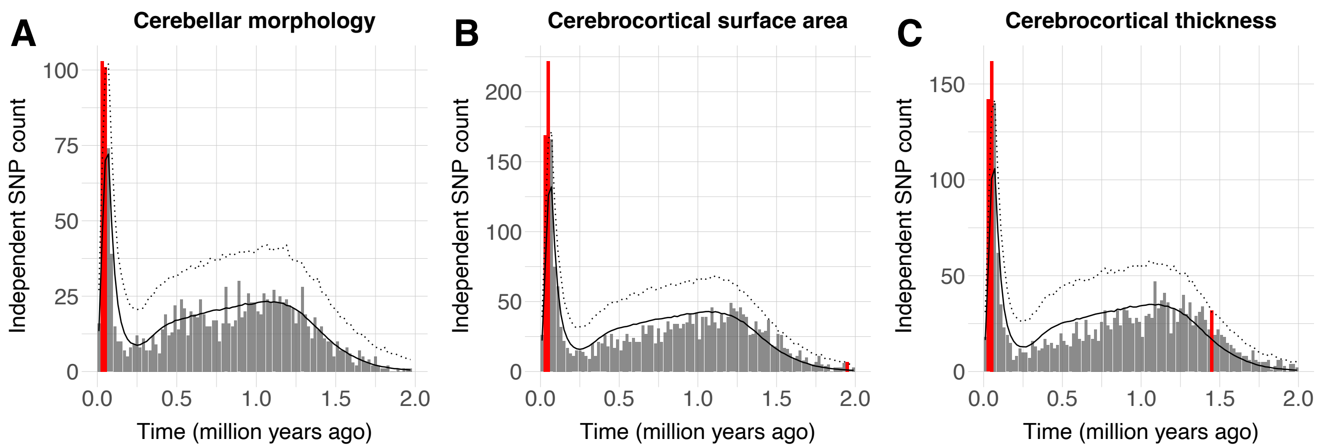


***Supplementary Figure 13: Independent SNPs associated with cerebellar and cerebrocortical morphology are enriched for evolutionary recent mutations in the human genome. A:*** *Histogram of estimated SNP age (ranging from 0 to 2 million years, in bins on 20,000 years) for independent lead SNPS associated with A) cerebellar morphology (1936 SNPs), B) regional cerebrocortical surface area (3606 SNPs) and C) regional cerebrocortical thickness (2993 SNPs). The solid black and dotted lines denote the mean and upper 95th confidence interval (Bonferroni corrected across 100 time-bins) derived from a null model constructed from 10,000 of equally sized sets of SNPs randomly drawn from the Human Genome Dating Atlas of Variant Age (after matching these to brain-phenotype-related SNPs in terms of minor allele frequencies). Red bars denote time-bins of significant positive enrichment. See Supplementary Data 13 for full numerical results.*

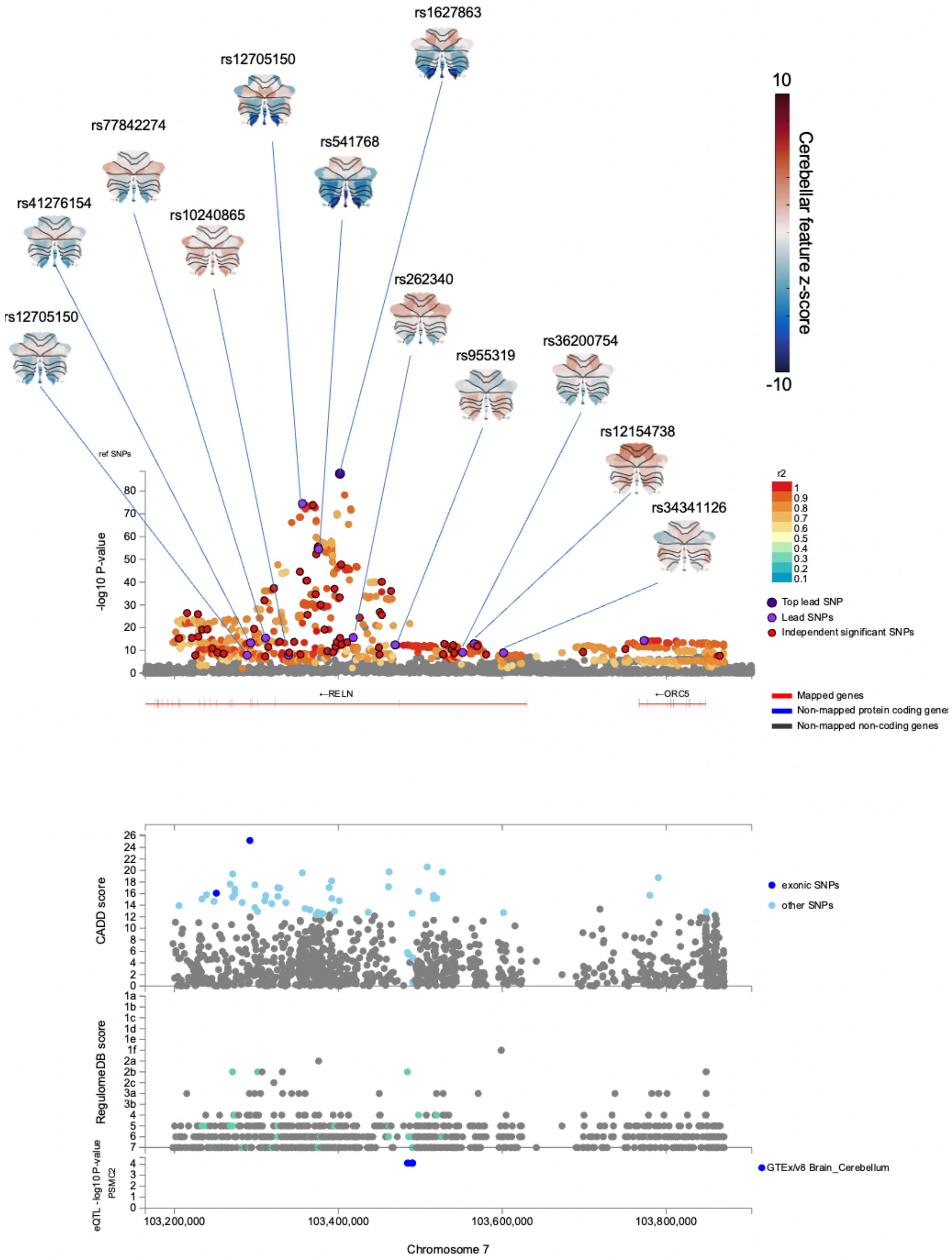


***Supplementary Figure 14:*** *Regional ´plot of locus 180, showing the 12 lead SNPs mapped to RELN and their associated z-score maps. While the strongest positive and negative z-scores overlapped cerebellar regions whose volume has previously associated with variations in the RELN gene (i.e., posterior and midline regions), these maps also demonstrate the distributed effects of RELN associated genetic variation across the cerebellar cortex*

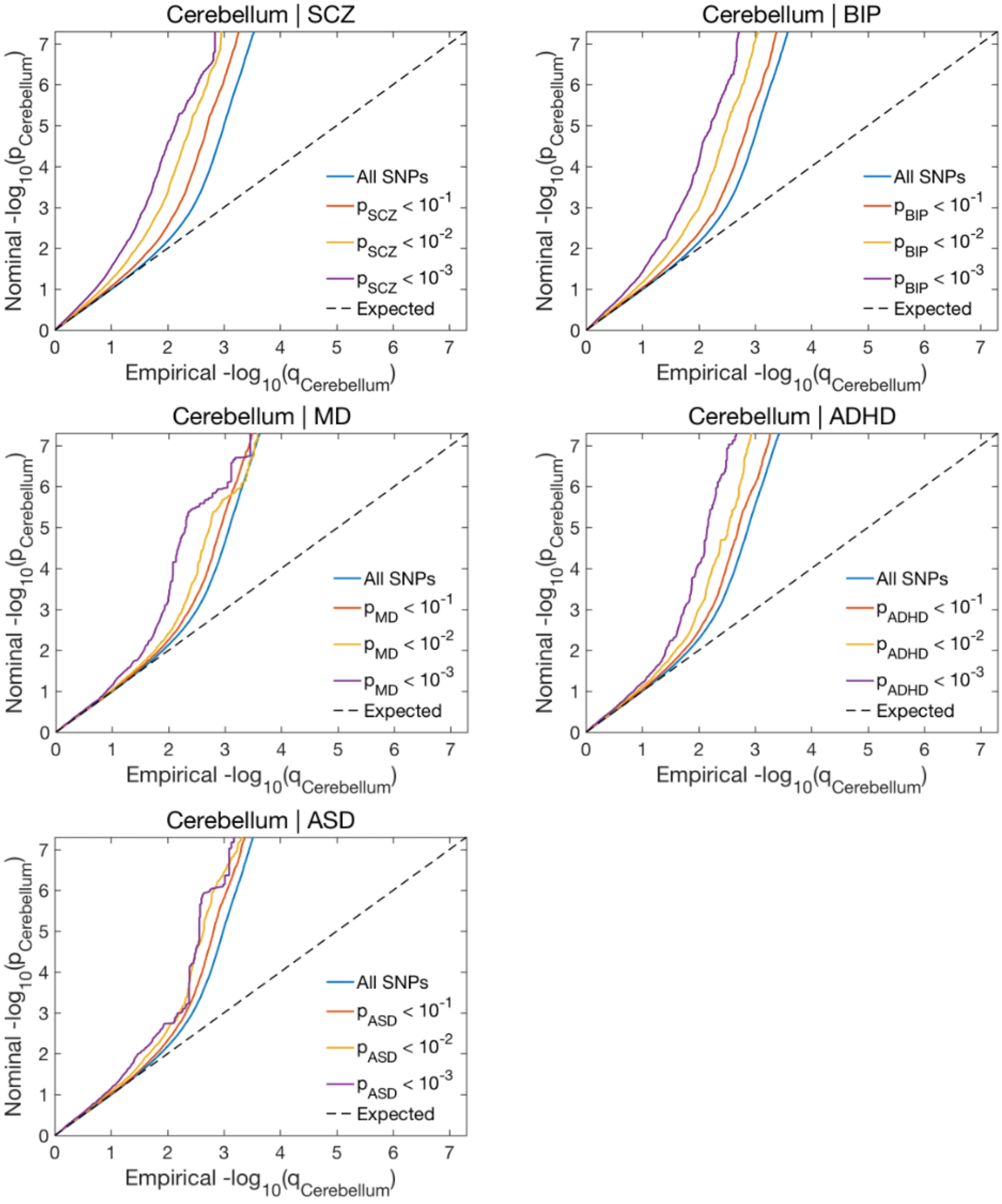


***Supplementary Figure 15:*** *QQ-plots from the conditional/conjunctional FDR analyses depicting the reverse association, i.e., enriched association with cerebellar morphology when conditioning on the association with psychiatric disorders.*
